# Parental perceptions of COVID-19-like illness in their children

**DOI:** 10.1101/2020.11.05.20226480

**Authors:** Ava Hodson, Lisa Woodland, Louise E Smith, G James Rubin

## Abstract

**Objectives:** To explore parents’ perceptions of COVID-19-like symptoms in their child and attitudes towards isolating from others in the household when unwell.

**Study Design:** Qualitative, semi-structured interviews.

**Methods:** 30 semi-structured telephone interviews with parents of children between 4 and 18 years.

**Results:** We found four themes relating to symptom attribution (‘normalising symptoms’, ‘err on the side of caution’, ‘experience of temperature’, ‘symptoms not normal for us’). In general, parents were more likely to attribute symptoms to COVID-19 if a temperature was present or the symptoms were perceived as ‘unusual’ for their family. Four themes relating to self-isolation (‘difficult to prevent contact with children’, ‘isolation would be no different to lockdown life’, ‘ability to get food and supplies’, ‘limited space’). Parents believed they would find isolation within the household difficult or impossible if they had dependent children, had limited space or could not shop for groceries.

**Conclusions:** The findings highlight complexities in symptom perception, attribution, and household isolation. We suggest that they can be overcome by a) providing better guidance on what symptoms require action, b) providing guidance as to how to prevent infection within the household, and c) by supporting families with grocery shopping through a potential second or third wave.

---

Children can spread certain illnesses readily at school and, in order to prevent outbreaks, are encouraged to stay at home when ill or until they are no longer infectious (1). How thoroughly children and parents adhere to the rules around sickness absence is unclear. In England, schools were closed nationally from 23^rd^ March 2020, to prevent the spread of COVID-19 (2) and re-opened to all children in September. There have been concerns that some parents may continue to send children to school when experiencing the symptoms of COVID-19, which include fever and cough (3).

Sending a child to school while symptomatic, rather than keeping them at home and arranging a test for COVID-19 as recommended by national guidance, may depend partly on how parents interpret their child’s symptoms (4) and partly on whether the parent is willing and able to keep the child off school. This in turn may require the parent to take time off work and affect the child’s education. However, little is known about factors that are at play in this decision-making process.

In this paper, we use data from interviews that explored parents’ perceptions of COVID-19-like symptoms in their child and attitudes towards isolating from others in the household when unwell.

Two interviewers conducted semi-structured telephone interviews lasting approximately 75 minutes between 15th and 21st April 2020 (n = 30, female = 20). All participants were aged 18 years or over and were the primary caregiver to a child who, from March 23rd, 2020 was not attending pre-school or school in England because of COVID-19. At the time of data collection, the recognised symptoms of COVID-19 were a new, continuous cough and fever. Loss of sense of smell or taste was added on May 18^th^, 2020 (5). At the time of interviewing, guidance stated that individuals who suspected they had symptoms should isolate as best they could from others in the household (6).

A broad discussion guide was used, covering psychological wellbeing, educational activity while at home and adherence to social distancing guidance. Results for these aspects of the interviews will be reported elsewhere. In this manuscript, we focus on responses relating to symptom perception, attribution, and how parents thought they would react to the presence of symptoms among their children. In our interview schedule, we asked parents whether their child had had “coronavirus or coronavirus symptoms, either a high temperature or new continuous cough?”. We also asked a set of questions about whether the parent or child would find it difficult to self-isolate and how they would cope with self-isolation.

Results were analysed using an inductive approach to thematic analysis (7). We found four themes relating to symptom attribution and four themes relating to self-isolation. These are described in Table 1 with supporting quotes.

**Table 1:**
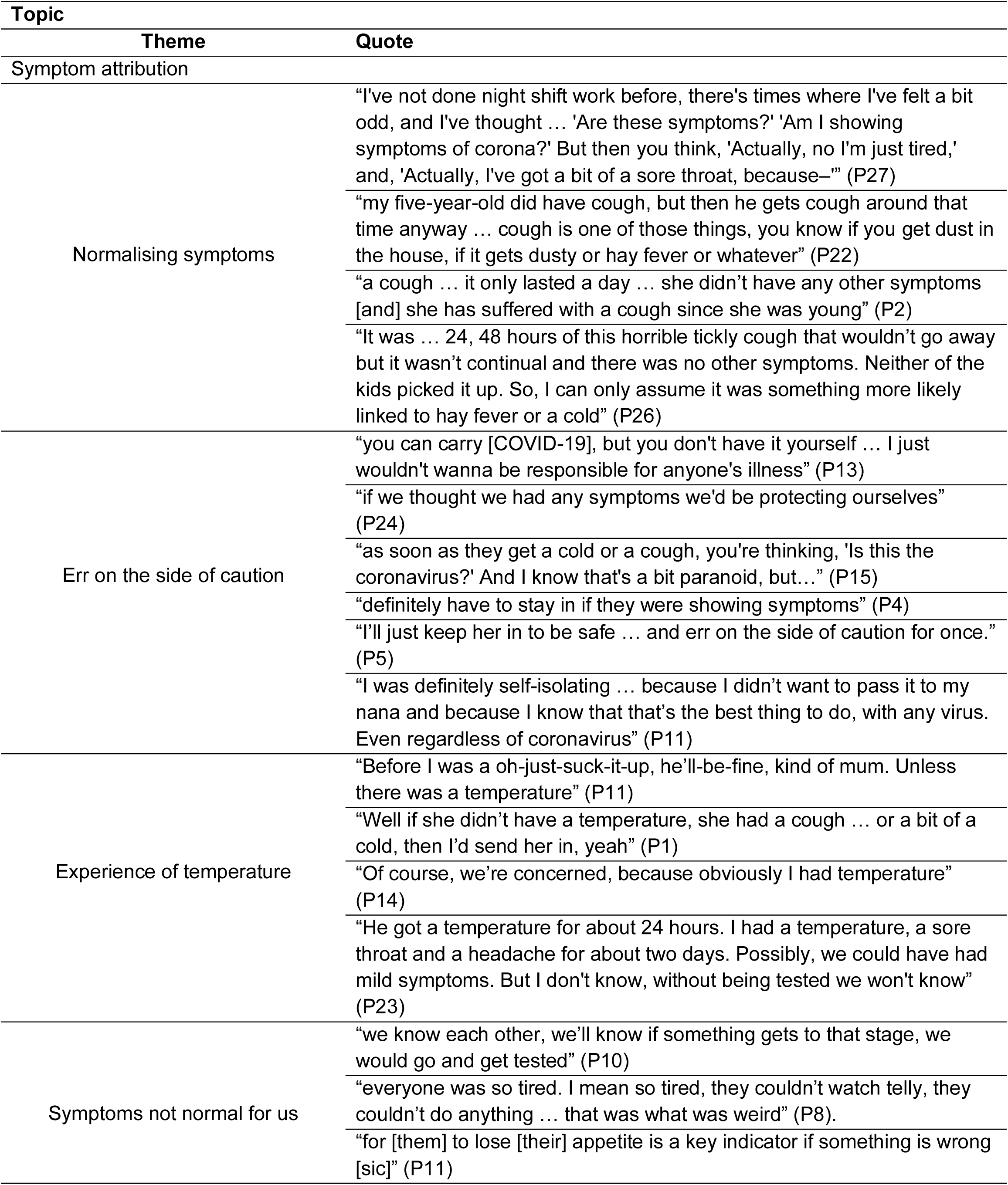
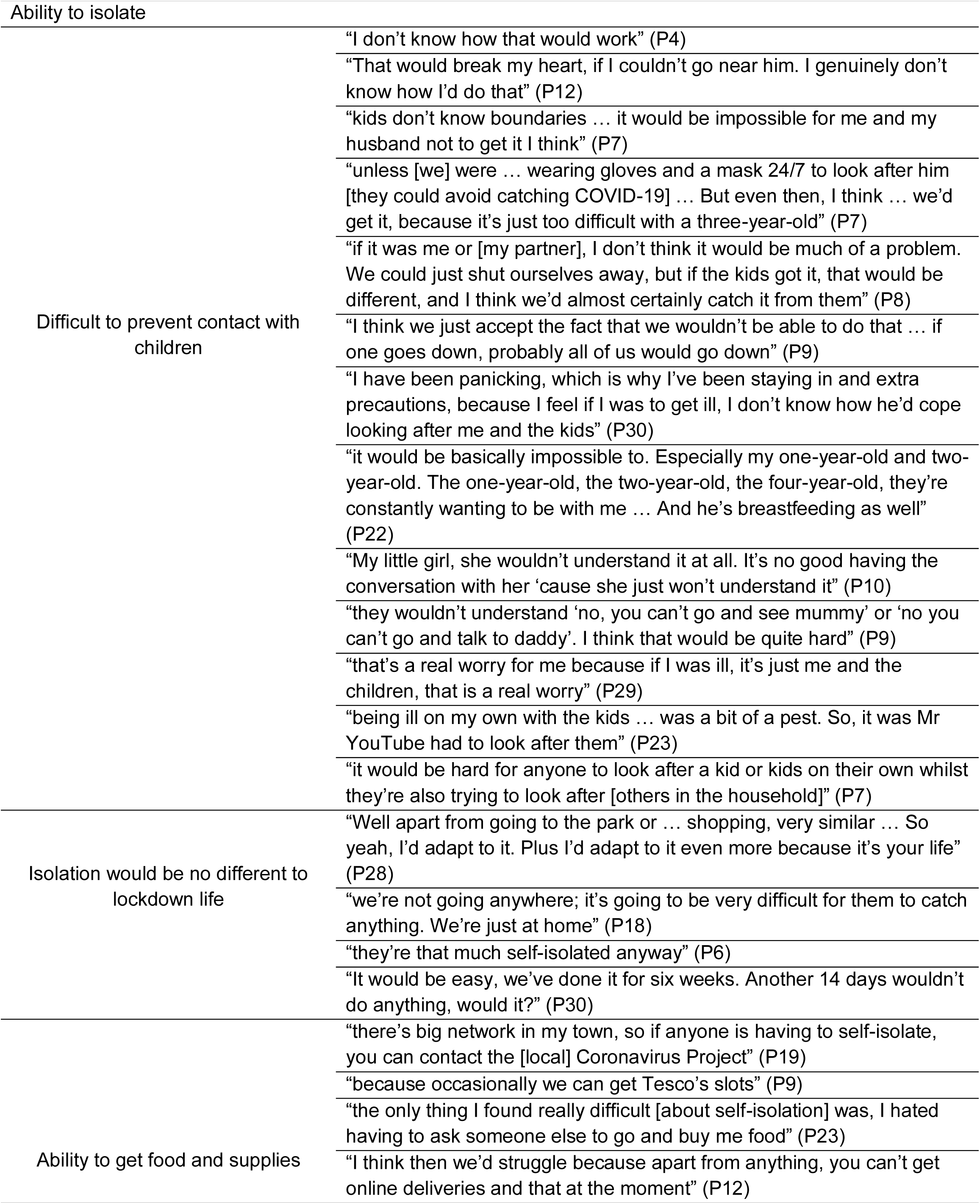
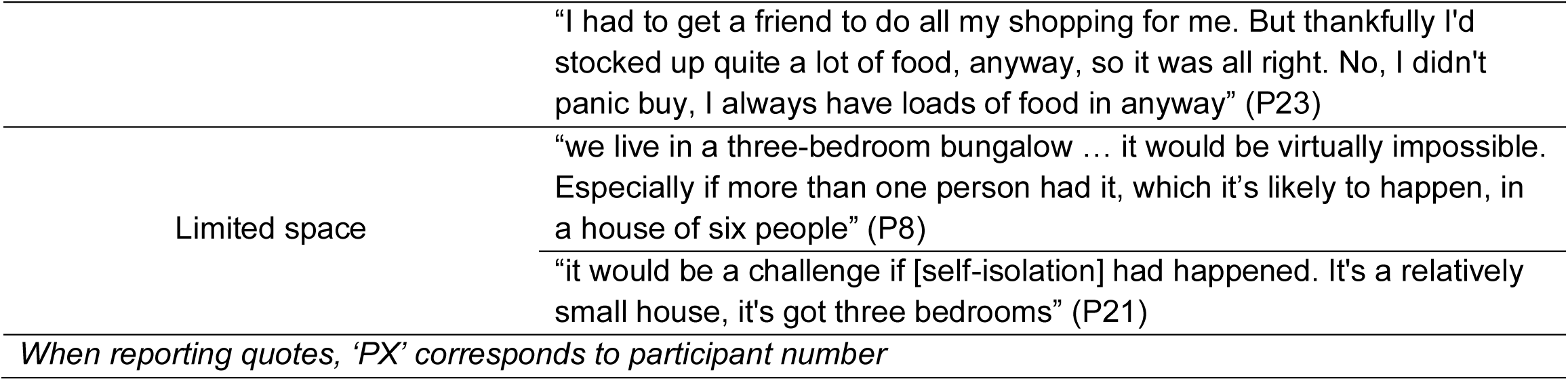
Supporting quotes for identified themes

In terms of symptom attribution, parents appeared to go through a process of finding the most likely reason for the experience of symptoms, discounting COVID-19 if a more likely explanation was apparent (“normalising symptoms”). Symptoms were often normalised in day-to-day terms that reduced the perceived risk and thus the intention to isolate. Particularly if symptoms were transitory or mild, this reduced worry and increased the likelihood of attribution to a non-COVID explanation. Conversely some participants expressed a view that “any symptoms” should be treated as if they were COVID-19 related (“err on the side of caution”). Given the context of the pandemic, they would isolate if they or a household member experienced either a cough or fever, to be on the safe side.

Some parents indicated that they would be more likely to attribute high temperature than a cough to COVID-19 (“experience of temperature”). While a cough could be put down to a sore throat or common cold, parents appeared more cautious about a temperature.

There was a sense among some participants that unexpected or unusual symptoms would be a particular cause for concern (“symptoms not normal for us”). Annual or seasonal experience of symptoms (e.g. hay fever) reduced parental concerns about whether symptoms were due to COVID-19 or not, as well as the experience of symptoms ‘normal’ to their household. Conversely, symptoms that were “weird” or unusual for the person were “a key indicator if something is wrong.”

Parents had varied beliefs about their ability to isolate from others in the home and particularly the difficulty of isolating from children (“difficult to prevent contact with children”). Across interviews, parents expressed that this was related to their child’s age and their understanding of the virus, i.e. younger children do not understand boundaries or reasons why they cannot be close to their parents. A common, fatalistic sentiment was that if one member of their household presented symptoms of COVID-19, then the whole household would catch it. Single parents relied on novel sources of care when they themselves became ill – for example, increased dependence on online resources, such as gaming and YouTube videos. When having to isolate due to symptoms, one parent stated they sent their child to their grandparent’s house so that they could get sufficient rest to recover.

At the time of data collection, people could only leave home for very limited reasons (for example, shopping as infrequently as possible and a daily walk or exercise). Some parents noted that it would be easy to isolate the entire household as it would be no different to how they were already living during lockdown (“isolation would be no different to lockdown life”).

Access to additional help or available resources was identified as impacting parents’ ability to isolate. It would appear that ability to self-isolate was facilitated by connectedness to other members of the community and access to local shops (“ability to get food and supplies”); lack of this may make parents believe that they are not able to isolate. Some parents identified that the size of the home would be an additional challenge during self-isolation (“limited space”).

Although rules and context have changed since the data were collected early in the pandemic, the findings highlight several key areas worthy of further exploration and quantification. We believe that there are two main implications.

First, parents’ perceptions of whether a given symptom is a possible indicator of COVID-19 do not match the official guidance. Symptoms are often not attributed to COVID-19 unless a temperature is present. Data from a national UK study suggests that amongst under 18s, 48% of those who tested positive for COVID-19 reported having a temperature in the first 7 days of the illness (8). Since parents who identified “not normal” symptoms or a temperature in themselves or their child were more inclined to attribute them to COVID-19, communications may benefit from highlighting that the presence of even one of the identified symptoms – i.e. a cough alone – necessitates self-isolation or request of a test, even if that symptom is mild (6).

Second, isolation is seen as difficult by many parents. Given the guidance at the time of interviewing, parents ideally should have isolated from their children to the best of their ability. Many parents identified that this would be difficult or “impossible”. Our findings also suggest that larger families and those living in smaller homes may find it particularly difficult to isolate. Indeed, research suggests that households with dependent children are less likely to adhere to self-isolation than those without children (9). Those who perceived household isolation to be easier were those who said it would be no different to their life in lockdown and would isolate even if they were not sure their or their child’s symptoms were due to COVID-19. Parents who reported that they could rely on someone for help with food shopping were more likely to think they could isolate. This is in line with previous research (10). We suggest that aiding families with grocery shopping may be a practical route to facilitating adherence to isolation. While it might be impractical for supermarkets to do this, grassroots organisations played a key role in the first wave and could mobilise again in the second wave.

## Data Availability

All data referred to in the manuscript can be made available if necessary.

